# Validated ICP-MS method for measurement of plasma and intracellular antimony concentrations applied to pharmacokinetics of meglumine antimoniate

**DOI:** 10.1101/2020.09.15.20194647

**Authors:** Diana J. Garay-Baquero, David E. Rebellon-Sánchez, Miguel D. Prieto, Lina Giraldo-Parra, Adriana Navas, Sheryl Atkinson, Stuart McDougall, Maria Adelaida Gómez

## Abstract

**Aim:** A high-throughput method using inductively coupled plasma mass spectrometry (ICP-MS) was developed and validated for the quantitative analysis of antimony in human plasma and peripheral blood mononuclear cells (PBMCs) from patients with cutaneous leishmaniasis undergoing treatment with meglumine antimoniate.

**Methods:** For this study, antimony was digested in clinical samples with 1% TMAH / 1% EDTA and indium was used as internal standard. Calibration curves for antimony, over the range of 25 to 10000 ng/mL were fitted to a linear model using a weighting of 1/concentration^2^. Accuracy, precision and stability were evaluated.

**Results:** Taking the lower limit of quantitation (LLOQ) to be the lowest validation concentration with precision and accuracy within 20% (25% at the LLOQ), the current assay was successfully validated from 25 to 10000 ng/mL for antimony in human plasma and PBMCs. Dilution studies demonstrated that concentrations up to 100000 ng/mL of antimony in plasma were reliably analyzed when diluted into the calibration range.

**Conclusion:** This protocol will serve as a baseline for future analytical designs, aiming to provide a reference method to allow inter-study comparisons.

What is already known about this subject
- Antimonial drugs are the mainstay treatment for cutaneous leishmaniasis of which systemically administered pentavalent antimonials (SbV) are widely used, however the pharmacokinetics (PK) of these drugs at the site of action is unknown.
- A wide range of analytical strategies have been used to quantify antimony in biological samples and atomic absorption spectroscopy is the most employed technique, however, no standardized methods for determination of intracellular concentrations of antimony were available.
- Relationships between plasma and intracellular drug concentrations remain unknown for most antiparasitic drugs, and PK studies rely on plasma drug concentrations assuming these act as surrogates of intracellular concentrations.

What this study adds
- We have developed and validated a reproducible and accurate ICP-MS method for the quantification of total antimony in human plasma and peripheral blood mononuclear cells (PBMCs) in accordance with the European Bioanalysis Forum (EBF) recommendations.
- This method was successfully used to compare pharmacokinetic curves of antimony in plasma and intracellular compartments, in samples collected from patients undergoing treatment for cutaneous leishmaniasis with meglumine antimoniate.

## INTRODUCTION

Leishmaniasis is a group of diseases caused by protozoan parasites of the genus *Leishmania*, which can cause cutaneous, mucosal or visceral manifestations depending on the infecting species and the host immune status ^1,2^. According to the most recent report from the World Health Organization (WHO), leishmaniasis is endemic across all continents. Despite an elevated proportion of suspected underreporting, 700000 to over 1200000 new cases are estimated to occur annually ^2,3^.

*Leishmania* is an intracellular parasite that invades and replicates predominantly within phagocytic cells (primarily macrophages), providing a “protected” environment against host defenses and a barrier to drug exposure. Antimonial drugs are the mainstay treatment for cutaneous leishmaniasis of which systemically administered pentavalent antimonials (Sb^V^) are widely used in the Americas and as intradermal injections in the Old World ^4^. Sodium stibogluconate (Pentostam®) and meglumine antimoniate (Glucantime®) are the synthetic compounds used for pentavalent antimonial therapy. Their efficacy is highly variable, according to the infecting species, parasite subpopulations, host responses and therapeutic regimens ^5-7^. Antileishmanial drugs need to be distributed to the parasite-targeted tissues (primarily skin, liver, spleen and bone marrow depending on the infecting species), internalized into host cells and distributed to the phagolysosomal compartment where the parasite resides ^8^. Although antimonials have been used over decades, the pharmacokinetics (PK) and pharmacodynamics (PD) of these drugs are not completely understood ^9,10^. Few studies have described antimonial PK in the context of human leishmaniasis ^11,12^ and the plasma and intracellular PK parameters involved in the therapeutic response remain unknown.

Atomic absorption spectrometry is the most common method used to quantitatively determine antimony levels in biological specimens such as blood and urine from patients undergoing antimonial therapy in pharmacokinetic studies ^11,13^. Although, a validated analytical method for quantification of arsenic and antimony in liposomes was previously developed using inductively coupled plasma-optical emission spectrometry ^14^, there is not a validated method available for intracellular antimony determination. Inductively coupled plasma mass spectrometry (ICP-MS) is an ultrasensitive and selective method that allows quantification levels in the order of parts per trillion, with a wide dynamic range of nine orders of magnitude and high throughput ^15^. Interferences are relatively low and matrix effects can be minimized by using internal standard controls ^16,17^. ICP-MS has been widely used in pharmacokinetic studies in humans of metal-based formulations such as arsenic, gadolinium, and some antineoplastic drugs ^18-22^. Its convenience and reduced time-consumption make this technique a powerful tool for quantitative determination of antimony in clinical samples at intracellular levels.

In this study, we present a scientifically validated analytical protocol for determination of total antimony in human plasma and peripheral blood mononuclear cells from patients with cutaneous leishmaniasis undergoing treatment with meglumine antimoniate (Figure 1).

**Figure 1.**
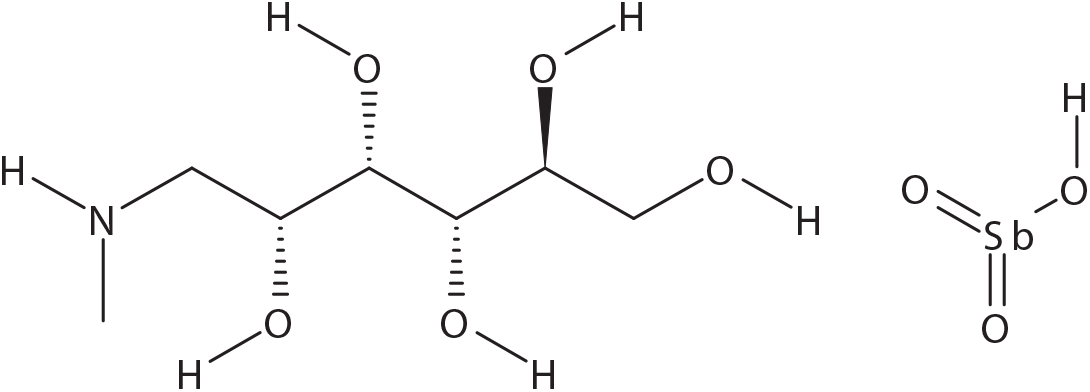
Meglumine antimonate chemical structure.

## METHODS

### Reagents

ICP grade antimony (Batches 884136D and BCBP51619V) and indium (Batch 833920k) were obtained from Alfa Aesar (Heysham, UK) and Sigma Aldrich (Buchs, Switzerland). Electronic grade tetramethylammonium Hydroxide (TMAH) was purchased from Alfa Aesar and ethylenediaminetetraacetic acid (EDTA) from (Sigma). Human plasma for validation studies was obtained from Biochemed (Winchester, USA). High purity (>99.999%) argon gas (BOC Gases, UK) was used for ICP-MS analysis. ASTM Type I ultra-pure water (>18 MΩ) was provided in-house by an Elga Purelab-Ultra system (Veolia Water Technologies, UK).

### Clinical samples and study design

Plasma samples and peripheral blood mononuclear cells (PBMCs) were obtained from EDTA anticoagulated blood from patients with cutaneous leishmaniasis (CL, n=5). All CL patients had parasitological confirmation of infection and were recruited at Centro Internacional de Entrenamiento e Investigaciones médicas (CIDEIM) outpatient clinics in Cali, Colombia. CL patient samples were collected from a cohort study that evaluates the relationship between antileishmanials PK, immune response signatures and clinical responses (CIDEIM IRB Study code CIEIH-1258). The study was reviewed and monitored by CIDEIM ethics committee in accordance with national (resolution 8430, República de Colombia, Ministry of Health, 1993) and international (Declaration of Helsinki and amendments, World Medical Association, Fortaleza, Brasil, October 2013) guidelines.

Participants were treated with intramuscular meglumine antimoniate (Glucantime® [81 mg Sb/mL]; (Sanofi-Aventis) during 20 days with a dose of 20 mg/kg up to a maximum dose of 1620 mg of Sb(V), equivalent to a 20 mL injection. The median adherence to the indicated regime was 85% (75% 100%). Peripheral blood samples were collected during the last day of treatment at the following time-points: before dose and at 0.5, 1, 1.5, 2, 3, 5, 8, 12 and 24 hours after drug administration. In total, samples from 5 CL patients were included in this study for quantification (Table 1).

**Table 1.**
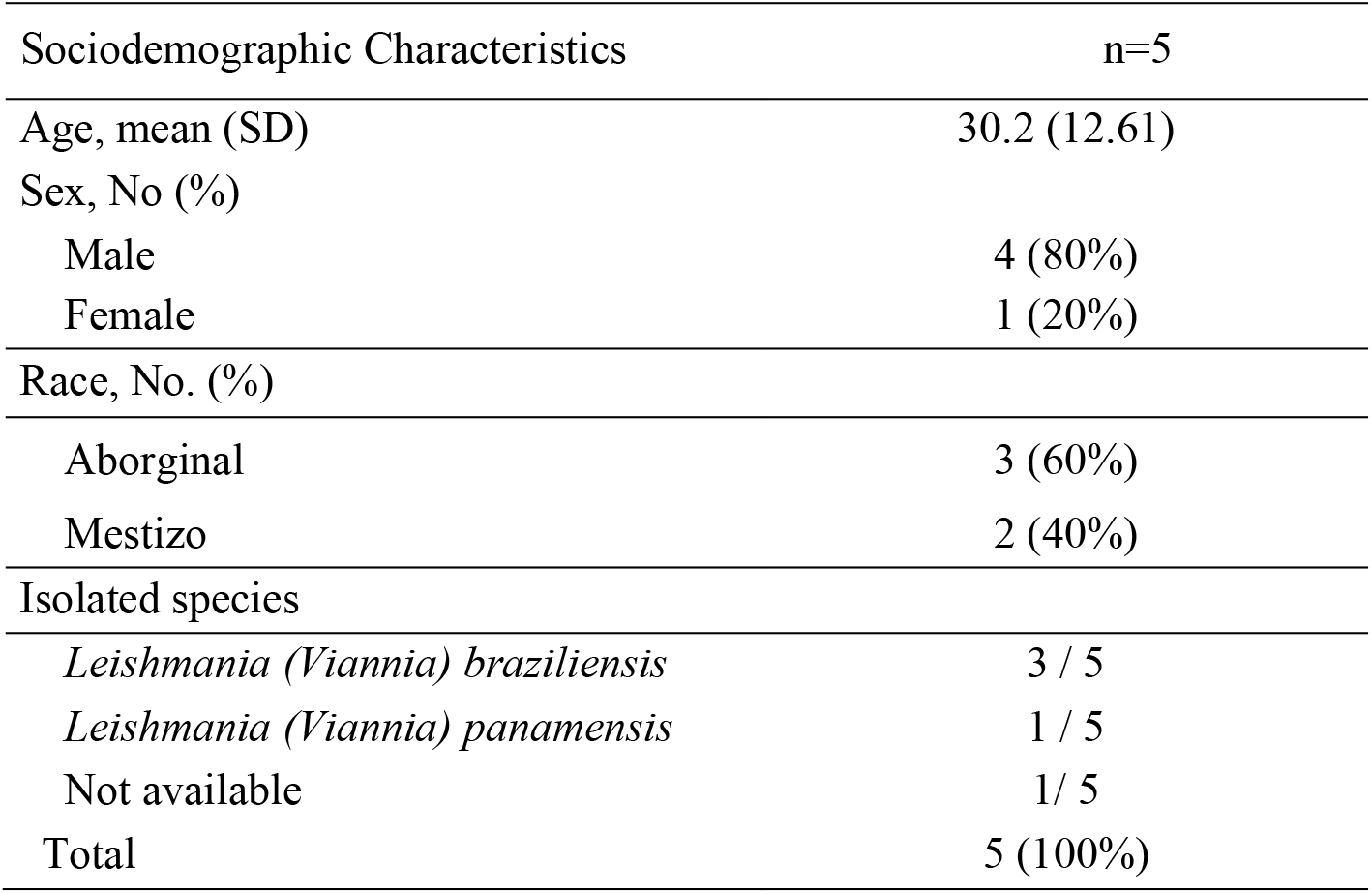
Clinical characteristics of CL patients

| Sociodemographic Characteristics | n=5 |
| --- | --- |
| Age, mean (SD) | 30.2 (12.61) |
| Sex, No (%) |  |
| Male | 4 (80%) |
| Female | 1 (20%) |
| Race, No. (%) |  |
| Aboriginal | 3 (60%) |
| Mestizo | 2 (40%) |
| Isolated species |  |
| *Leishmania (Viannia) braziliensis* | 3 / 5 |
| *Leishmania (Viannia) panamensis* | 1 / 5 |
| Not available | 1/ 5 |
| Total | 5 (100%) |

### Plasma procurement and PBMC isolation

Venous blood was collected in EDTA vacutainer tubes and plasma was prepared by centrifuging 10 mL of blood at 1300 x g for 10 minutes at room temperature. An aliquot of 3 mL of plasma was transferred to a fresh tube and centrifuged again under the same conditions. The plasma was transferred to a new tube and stored at −80°C. Remaining plasma and blood cells were gently mixed with PBS to complete 20mL final volume. Blood/PBS mix was carefully layered on top of 10 mL Ficoll-Hypaque 1077 gradient (Sigma) in a 50 mL centrifuge tube and centrifuged at 400 x g for 35min at room temperature (RT = 19 – 23 °C). The plasma layer was carefully removed, and the mononuclear cells layer was transferred to a separate centrifuge tube. Cells were washed with at least three volumes of PBS. Cells were subsequently centrifuged at 400 x g for 15 min at RT. Then, the supernatant was discarded and the pellet was resuspended in 200 µL of 25%TMAH and stored at approximately −80°C until use. This PBMC preparation is referred as PBMC matrix for method development.

### ICP-MS analysis

The analyses were performed in a dedicated ACDP (UK Advisory Committee on Dangerous Pathogens) Class II bio-facility and all sample processing and digestion was performed inside Class I HEPA filtered biosafety cabinets to minimize the risks of contamination. All elemental reference standard preparations (Sb and In) were performed in a separate and dedicated laboratory to further eliminate the potential for sample contamination

### Instrumentation and system suitability

Antimony and indium (internal standard), each have two stable isotopes: ^121^Sb and ^123^Sb, and ^113^In and ^115^In, respectively. Of these, ^121^Sb and ^115^In are most abundant and, therefore, were used to quantify antimony and indium, as internal standard, by ICP-MS.

All measurements were performed using an Agilent 7700x series ICP-MS, and used to quantify antimony (mass 121) relative to the indium internal standard (mass 115), following infusion of the sample into the ICP-MS via an integrated sample introduction system (ISIS) and micromist nebulizer. The operating system was the ICP MassHunter, version B.01.03 (Agilent). Operating parameters are listed in Table 2. The regression and quantitation procedures were performed using Watson LIMS version 7.5 SP1 (Thermo Fisher Scientific, USA).

**Table 2.** Agilent 7700x operating parameters

| ICP-MS | Setting |
| --- | --- |
| RF Power | 1550W |
| # of points per peak | 3 |
| Replicates | 3 |
| Integration time/mass | 0.09 s (<sup>115</sup>In); 0.09 s (<sup>121</sup>Sb) |
| Total acquisition time | 1.98 s |
| Nebulizer/carrier gas flow | 1.05 L/min |
| Dilution mode | OFF |
| Integrated Sample Introduction System (ISIS-DS) | Setting |
| Load time | 8 s |
| Load speed | 0.45 rps |
| Probe rinse time | 90 s |
| Post rinse time | 30 s |
| Post rinse speed | 0.5 rps |
| Loop tubing ID | 2.29 mm |

Prior to initiating any run on the system, an ICP-MS tune test was performed by analyzing a calibration mix containing cerium, cobalt, lithium, magnesium, thallium and yttrium (10 ppm, Agilent) in Tune mode. The raw counts should exceed 1000 units for each element and the variability (% relative standard deviation, %RSD) should be under 10% for the system to be classified as suitable. Additionally, a system suitability test (SST) was executed by running two antimony dilutions; the low SST solution was 0.5 ng/mL and the high SST solution was 200 ng/mL. These dilutions were prepared using the indium internal standard solution (InISS) as matrix. InISS was a dilution at 2 ng/mL of Indium in 1%TMAH / 1%EDTA. The blank was a dilution of 2% human plasma matrix in 1%TMAH / 1%EDTA.The raw antimony count of the blank must be ≤ 25% of the low SST raw count.

### Preparation of calibration and quality control standards

Calibration standards were prepared by mixing an aliquot of the commercial antimony stock solution with purified water to yield eight calibration standards 2.5, 4, 25, 50, 250, 500, 725, 1000 µg/mL. Similarly, a set of five quality control (QC) standards were prepared separately with purified water to produce 2.5, 5, 50, 750, 10000 µg/mL concentrations.

### Preparation of calibration samples

Calibration stock standards were mixed with filtered control human plasma in a rotary mixer for at least 30 min at 30 rpm to yield nominal antimony concentrations of 25, 50, 250, 500, 2500, 5000, 7500 and 10000 ng/mL. Calibration samples were either used fresh or stored frozen at either −20 or 80 ± 10°C for no longer than 56 days. Calibration samples used to determine stability were freshly prepared.

### Preparation of validation samples

Plasma validation samples were prepared by mixing QC standard solutions with human plasma in a rotary mixer for at least 30 min at 30 rpm to give nominal antimony concentrations of 25 (QCLLOQ), 50 (QCL), 500 (QCM), 7500 (QCH) and 100000 ng/mL (DQC). Validation samples were stored frozen at either −20 or −80 ± 10°C for no longer than 56 days.

PBMC validation samples were prepared by mixing an aliquot (100 μL) of the plasma validation samples QCLLOQ, QCL, QCM, and QCH with and an aliquot of control PBMC matrix (100 μL) at the time of sample digestion.

### Antimony digest preparation

Analytical samples were thawed at room temperature and vortex mixed. For plasma analysis, an aliquot of each plasma sample (100 µL) was mixed with 4.90 mL of InISS in a rotary mixer for 30 minutes at room temperature. For PBMC analysis, an aliquot of each PMBC sample (100 μL PBMC matrix and 100 μL plasma validation sample) was added to 4.80 mL of InISS and mixed for 30 min. An aliquot of each processed sample was then analyzed by ICP-MS together with quality control samples and a multilevel calibration.

Intracellular antimony concentration was calculated by dividing the total estimated antimony content in the cell pellet by the total cell volume (number of PBMCs per pellet × 283 fL, where 283 fL is the average volume of a single PBMC ^23^)

### Dilution

A 1:10 dilution of the samples was prepared by adding 0.5 mL of each digested sample to 4.5 mL of InISS. Samples were vortex mixed for approximately 10 seconds, tumbled for at least 30 minutes on rotary mixer and then transferred to a sample rack for analysis by ICP-MS.

## RESULTS

### Assay specificity and calibration

The spectrographic interference from antimony on the indium response was within acceptance criteria (≤ 5.00% of the mean peak area of indium matrix blank response in the run). Similarly, the spectrographic interference from indium on the antimony response was within acceptance criteria (≤ 25.0% of the mean peak area of the LLOQ of antimony response in the run. A linear model method using a weighting of 1/concentration^2^ was selected for calibration of antimony in human plasma and a representative calibration curve is presented in Figure 2.

**Figure 2.**
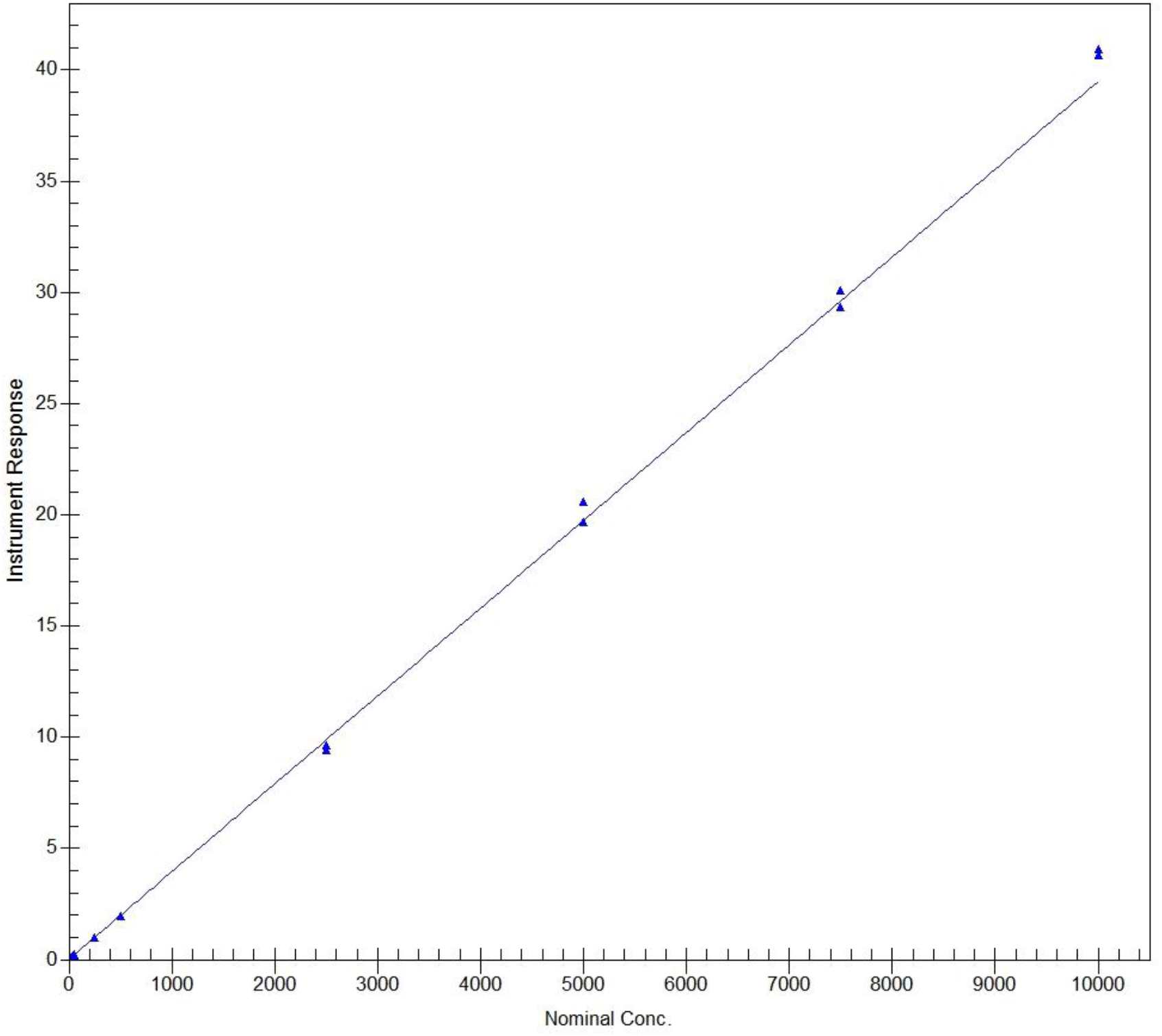
Representative calibration curve for antimony in human plasma. Calibration standards for antimony (25, 50, 250, 500, 2500, 5000, 7500 and 10000 ng/mL) were quantified using ICP-MS and calibration was fitted with a linear model based on weighting of 1/concentration2. This calibration was obtained from the analytical run 1 and processed with the software Watson LIMS version 7.5 SP1.

The calibration curve parameters obtained throughout the validation study are listed in Table 3.

**Table 3.** Calibration curve regression parameters for antimony

| Run day | Analytical run number | Slope | Intercept | R-Squared | LLOQ | LOQ |
| --- | --- | --- | --- | --- | --- | --- |
| 1 | 1 | 0.0039481 | 0.0089183 | 0.9966 | 25 | 10000 |
| 1 | 2 | 0.0038862 | 0.0082196 | 0.9979 | 25 | 10000 |
| 40 | 3 | 0.0039172 | 0.018297 | 0.9969 | 25 | 10000 |
| 68 | 4 | 0.0032206 | 0.0037602 | 0.9981 | 25 | 10000 |
| 68 | 5 | 0.0031721 | 0.0065024 | 0.9981 | 25 | 10000 |
| 75 | 6 | 0.0035459 | 0.0058869 | 0.9964 | 25 | 10000 |
| 132 | 7 | 0.00386 | 0.022976 | 0.9953 | 25 | 10000 |
| Mean |  | 0.0036500 | 0.0106515 | 0.9970 |  |  |
| SD |  | 0.0003376 | 0.0071493 | 0.0011 |  |  |
| %CV |  | 9.25 | 67.12 | 0.11 |  |  |

Quality control samples, prepared in plasma at antimony concentrations 50, 500, 7500 ng/mL were used to demonstrate the performance of the analytical assay. The relative bias of the back-calculated concentrations of antimony within the acceptance criteria ranged between −2.60% and 3.51% for 7 analytical runs ran over 132 days (Table 4).

**Table 4.** Back-calculated calibration standards for antimony in human plasma

| Analytical run number | 25.0 ng/mL | 50.0 ng/mL | 250 ng/mL | 500 ng/mL | 2500 ng/mL | 5000 ng/mL | 7500 ng/mL | 10000 ng/mL |
| --- | --- | --- | --- | --- | --- | --- | --- | --- |
| 1 | 21.9 | 48.5 | 246 | 494 | 2390 | 4980 | 7420 | 10300 |
|  | 28.0 | 52.2 | 250 | 489 | 2440 | 5210 | 7630 | 10400 |
| 2 | 23.8 | 50.2 | 238 | 501 | 2470 | 4990 | 7370 | 10600 |
|  | 27.1 | 47.2 | 243 | 485 | 2480 | 5180 | 7780 | 10200 |
| 3 | 23.8 | 45.3 | 247 | 512 | 2420 | 5260 | 7430 | 10600 |
|  | 27.6 | 49.0 | 245 | 514 | 2400 | 5180 | 7620 | 9580 |
| 4 | 26.0 | 49.7 | 252 | 504 | 2510 | 5350 | 7580 | 10700 |
|  | 25.1 | 46.3 | 240 | 483 | 2460 | 5050 | 7120 | 9970 |
| 5 | 24.3 | 47.2 | 246 | 486 | 2500 | 5150 | 7460 | 10400 |
|  | 26.6 | 48.7 | 261 | 512 | 2430 | 4880 | 7130 | 10600 |
| 6 | 23.4 | 49.5 | 249 | 463 | 2260 | 5350 | 7630 | 9410 |
|  | 25.7 | 54.0 | 258 | 508 | 2500 | 5200 | 7680 | *13400 |
| 7 | 23.6 | 50.2 | 254 | 503 | 2390 | 4920 | 7180 | 10900 |
|  | 27.8 | 44.2 | 241 | 501 | 2490 | 4830 | 7670 | 10900 |
| Mean | 25.3 | 48.7 | 248 | 497 | 2439 | 5109 | 7479 | 10351 |
| S.D. | 1.92 | 2.61 | 6.69 | 14.3 | 66.2 | 169 | 215 | 462 |
| %CV | 7.59 | 5.36 | 2.70 | 2.88 | 2.71 | 3.31 | 2.87 | 4.46 |
| %Bias | 1.20 | -2.60 | -0.80 | -0.60 | -2.44 | 2.18 | -0.28 | 3.51 |
| n | 14 | 14 | 14 | 14 | 14 | 14 | 14 | 13 |

Reason for exclusion
\* >20% of acceptance criteria, not in statistics

Quality control (QC) data was obtained during the validation process. Quality control samples at low (50 ng/mL), medium (500 ng/mL) and high (750 ng/mL) concentration were evaluated by duplicate over analytical runs 2 to 6 (n=10). The mean antimony concentration for the low concentration control was 50 ng/mL (SD=3.89; %CV=7.78; %Bias=0.00); 494 ng/mL (SD=9.91; %CV=2.01; %Bias=-1.20) for the medium concentration control and 7520 ng/mL (SD=189; %CV=2.51; %Bias=0.27) for the high concentration control sample. The impact of carry-over was assessed at the start and the end of every run containing a calibration curve. The mean percentage carry-over was 10.6 % and within the acceptance criteria (≤ 25.0% difference).

### Assay variability

Accuracy and within-run precision for the validation of the antimony assay are summarized in Table 5. The accuracy ranged from −1.20 to 1.20% of nominal values. At all concentration levels of antimony, the ranges of accuracy and precision were within established acceptance criteria.

**Table 5.** Accuracy and precision of antimony in human plasma

| Analytical<br>run number | 25.0 ng/mL | 50.0 ng/mL | 500 ng/mL | 7500 ng/mL |
| --- | --- | --- | --- | --- |
| 1 | 24.8 | 55.1 | 486 | 7420 |
|  | 26.1 | 48.6 | 486 | 7600 |
|  | 26.5 | 47.9 | 491 | 7570 |
|  | 23.2 | 46 | 510 | 7630 |
|  | 24.2 | 50.6 | 495 | 7610 |
|  | 26.1 | 50.6 | 498 | 7680 |
| Mean | 25.2 | 49.8 | 494.0 | 7585 |
| S.D. | 1.30 | 3.13 | 9.05 | 88.71 |
| %CV | 5.16 | 6.29 | 1.83 | 1.17 |
| %Bias | 0.80 | -0.40 | -1.20 | 1.13 |
| n | 6 | 6 | 6 | 6 |

Accuracy and within-run precision for the validation of the antimony assay in PBMC are summarized in Table 6. The accuracy ranged from −12.2 to 20.8% of nominal values. At all concentration levels of antimony, the ranges of accuracy and precision were within established acceptance criteria

**Table 6.** Accuracy and precision of antimony in human PBMCs

| Analytical<br>run number | 25.0 ng/mL | 50.0 ng/mL | 500 ng/mL | 7500 ng/mL |
| --- | --- | --- | --- | --- |
| 7 | 27.6 | 43.8 | 437 | 6390 |
|  | 31.3 | 42.3 | 454 | 6440 |
|  | 34.3 | 45 | 446 | 7050 |
|  | 28.4 | 44.2 | 420 | 7310 |
|  | 29.3 | 44 | 454 | 7450 |
| Mean | 30.2 | 43.9 | 442 | 6928 |
| S.D. | 2.68 | 3.13 | 9.05 | 88.70 |
| %CV | 8.88 | 7.14 | 2.05 | 1.28 |
| %Bias | 20.7 | -12.3 | -11.6 | -7.63 |
| n | 5 | 5 | 5 | 5 |

### Antimony stability

The stability of antimony in plasma was established following storage at room temperature for 24 hours using plasma samples spiked with 50 ng/mL or 7500 ng/mL of antimony standards defined as low and high concentration quantification controls, respectively. Six replicates were analyzed for each concentration and condition. No significant changes in the plasma antimony concentrations were observed after this storage period. The mean concentration for the low concentration control was 50.4 ng/mL (SD=4.28; %CV=8.49; %Bias=0.80) and 7550 ng/mL (SD=123; %CV=1.63; %Bias=0.67) for the high concentration control.

Additionally, stability of antimony in plasma was assessed following four freeze/thaw cycles at −20°C and −80°C ± 10°C utilizing plasma samples spiked with 50 ng/mL or 7500 ng/mL of antimony standard. Six replicates were analyzed for each concentration and condition and the accuracy and precision for quantitation of antimony under these conditions was evaluated. Subsequent to four freeze/thaw cycles at −20°C the mean concentration for the low concentration control was 48.3 ng/mL (SD=2.09; %CV=4.33; %Bias=-3.40) and 7290ng/mL (SD=314; %CV=4.31; %Bias=-2.80) for the high concentration control. At −80ºC the mean concentration for the low concentration control was 46.3 ng/mL (SD=1.18; %CV=2.55; %Bias=-7.40) and 7530 ng/mL (SD=107; %CV=1.42; %Bias=0.40).

Stability of antimony in plasma was established following storage at approximately −20°C and −80°C ± 10°C for 56 days. Six replicates were analyzed for each concentration and condition and the accuracy and precision for quantitation of antimony under these conditions was evaluated. Storage of plasma samples at −20ºC for 56 days resulted in a mean concentration for the low concentration control of 48.7 ng/mL (SD=1.66; %CV=3.41; %Bias=-2.60) and 7450ng/mL (SD=80.7; %CV=1.08; %Bias=-0.67) for the high concentration control. Storage at −80ºC for the same period resulted in a mean concentration for the low concentration control of 48.2ng/mL (SD=2.30; %CV=4.77; %Bias=-3.60) and 8150 ng/mL (SD=315; %CV=3.87; %Bias=8.67) for the high concentration control.

Stability of antimony in processed human plasma samples after storage at room temperature for 120 hours and 35 days at room temperature was also evaluated. Six replicates were analyzed for each concentration. Storage for 120 hours resulted in a mean concentration for the low concentration control of 52.0 ng/mL (SD=2.47; %CV=4.75; %Bias=4.00) and 7820ng/mL (SD=128; %CV=1.64; %Bias=4.27) for the high concentration control. Following storage of the processed samples for 35 days at room temperature resulted in a mean concentration for the low concentration control of 48.9 ng/mL (SD=1.14; %CV=2.33; %Bias=-2.20) and 7630 ng/mL (SD=158; %CV=2.07; %Bias=1.73) for the high concentration control.

### Influence of dilution on the quantitation of antimony in plasma

Plasma antimony levels can reach peak concentrations over 40 µg/mL following intramuscular administration of meglumine antimoniate ^11^, therefore plasma dilution is usually required for antimony quantification. The accuracy and precision results for quantitation of antimony following plasma sample dilution were evaluated over one analytical run and six replicates were measured. Plasma concentrations up to 100000 ng/mL were reliably analyzed when diluted 1:10 (v:v) into the calibration range of the assay. The mean quantified concentration of spiked plasma was 105000ng/mL (SD=2400; %CV=2.29; %Bias=5.00).

The accuracy and precision results for quantitation of antimony in digested dilutions prepared from plasma samples were also evaluated. Concentrations up to 100000 ng/mL were reliably analyzed when diluted 1:10 (v:v) into the calibration range of the assay. The mean concentration was 102000 (SD=816; %CV=0.80; %Bias=2.00, n=6).

### Antimony determination in clinical samples of patients undergoing antileishmanial treatment with meglumine antimoniate

The ICP-MS quantitative method validated in this study was applied to determine the plasma and intracellular levels of antimony in patients undergoing treatment for cutaneous leishmaniasis with meglumine antimoniate. Levels of antimony were measured at the end of the treatment (day 20) and evaluated up to 24 hours following drug administration. Figure 3A-E presents the plasma and intracellular (PBMCs) concentration-time curves of five adult patients.

**Figure 3.**
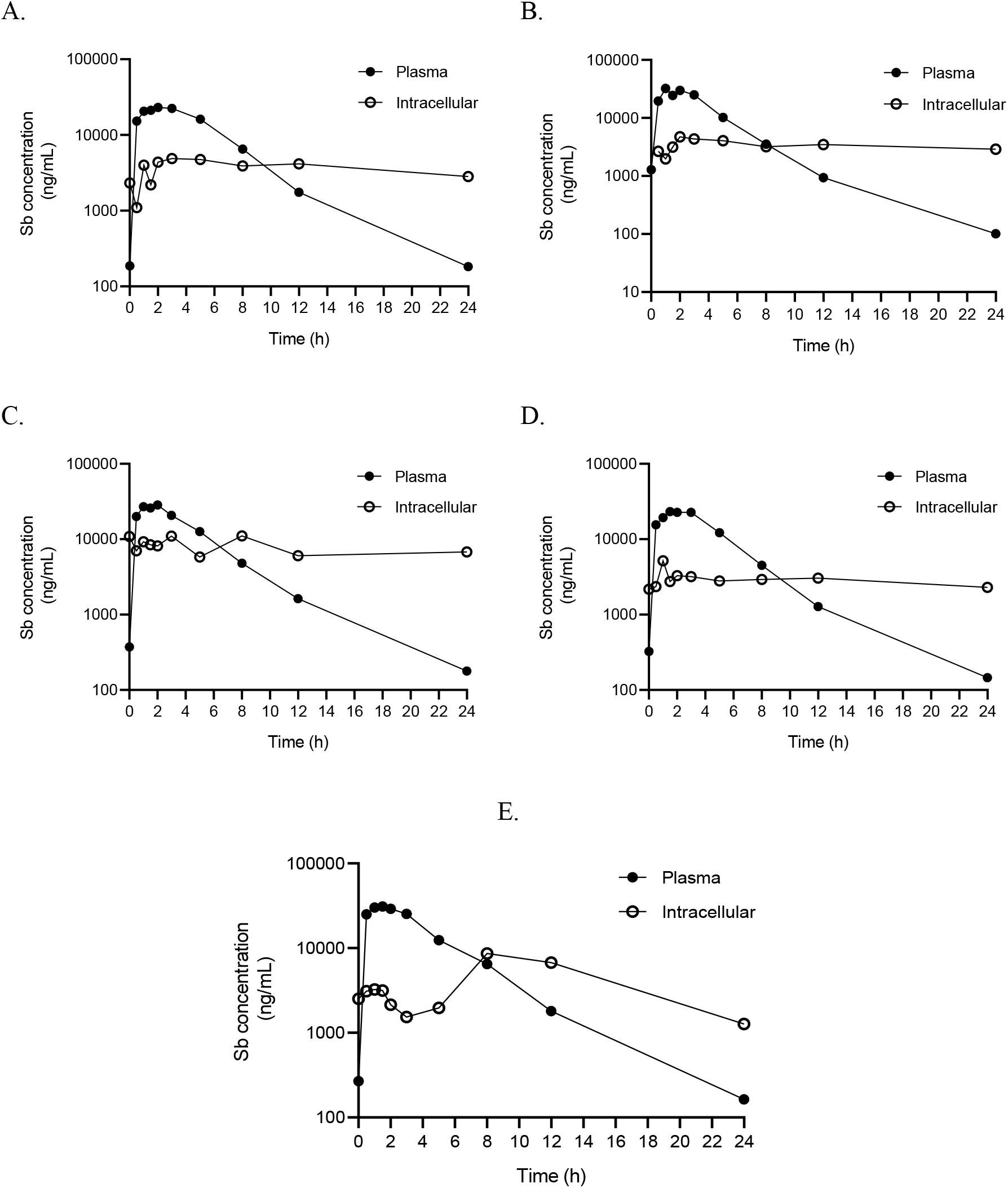
Antimony plasma and intracellular concentration-time curves. **(A-E)** Pharmacokinetic curves for antimony plasma and intracellular (PBMCs) levels. Samples were procured from five patients undergoing cutaneous antileishmanial therapy with meglumine antimoniate at the end of the treatment (day 20) and evaluated up to 24 hours following drug administration.

### Discussion

A wide range of analytical strategies have been used to quantify antimony in biological samples, and atomic absorption spectroscopy, using diverse atomization configurations, is the most employed technique ^24,25^. Previously, our group standardized and applied an electrothermal atomic absorption spectrometry method to quantify antimony in plasma and urine samples after administration of intramuscular Glucantime® ^11^. However, no standardized methods for determination of intracellular concentrations of antimony were available to date, which limited the study of pharmacokinetics of antimonial compounds at the site of drug action during antileishmanial chemotherapy.

We have developed and scientifically validated a reproducible and accurate ICP-MS method for the quantification of total antimony in human plasma and PBMCs in accordance with the European Bioanalysis Forum (EBF) recommendations. Antimony levels in clinical samples were quantitatively determined by ICP-MS following digestion in 1%TMAH / 1% EDTA, using indium as the internal standard. Accuracy and within-run precision were within acceptance limits. The maximum run size for this assay (assay robustness) was 240 injections per batch. Antimony was stable in human plasma at room temperature for at least 24 hours, at approximately −20°C or −80°C ± 10°C for at least 56 days and following at least four freeze/thaw cycles. In addition, processed samples were stable for at least 35 days at room temperature. Dilution evaluation demonstrated that concentrations up to 100000 ng/mL of antimony could be reliably analyzed when diluted into the calibration range. These data show that our method has adequate specifications to reliably perform pharmacokinetic studies of total plasmatic and intracellular antimony. Accuracy and precision for PBMC was not as high as in plasma, but met the predefined scientific validation assay acceptance criteria (+/-25% at LLOQ).

The main limitation of this method is that it does not allow Sb^III^ and Sb^V^ speciation. Antimony-based antileishmanial therapy is administrated as pentavalent antimony, which is rapidly absorbed reaching peak plasma concentration between 0.5h to 2h. However bio reduction to the trivalent form is required for the antileishmanial activity ^9^. It has been proposed that this redox process can occur in the host cell phagolysosome where the parasites reside, or in the parasitic cytosol ^26^. Sb^III^ induces an increase of intracellular Ca^2+^ and finally apoptosis. Thus, antileishmanial activity is highly dependent upon intracellular mechanisms. Although this method cannot discriminate antimony species, this is the first validated method available for intracellular quantification of antimony.

We generated pharmacokinetic curves of antimony in PBMCs from patients undergoing meglumine antimoniate antileishmanial treatment at last day of treatment and compared them to plasma concentration-time curves. Intracellular antimony concentration was over seven times higher than plasma before the last dose was administered showing intracellular accumulation over the course of the antileishmanial treatment. As expected, a faster peak concentration was reached in plasma compared to PBMCs after the last dose was administered. Intracellular drug quantification in chemotherapy against intracellular pathogens is key to dissect the mechanisms of susceptibility and therapeutic response, since adequate drug levels at the effect site, the cells in which the pathogen survives and replicates, determine the pharmacological activity. Despite this, the relationships between plasma and intracellular drug accumulation remain unknown for leishmaniasis and PK studies rely on plasma drug concentrations assuming these act as surrogates of intracellular concentrations ^27^. However, intracellular drug penetration can widely vary depending on host factors such as permeability, local metabolism and drug efflux/uptake transporters activity ^28^, which in turn, determine the level of exposure of intracellular pathogens to drugs. Additionally, multiple reports have shown a link between the host immune response and the antimonial therapy efficacy ^29^. Therefore, liking information about intracellular drug concentrations, host immune signals and therapy outcome can provide a powerful framework to optimize therapeutic regimens for *Leishmania*, as well as for other intracellular pathogens.

## Data Availability

All data are included within the main manuscript file

## Acknowledgements

We gratefully acknowledge the volunteers who participated in this study and members of the CIDEIM Clinical Unit. Authors declare no conflict of interest. The research reported in this publication was supported by Wellcome Trust 107595/Z/15/Z.

## Data availability

The data that support the findings of this study are presented within the manuscript. Clinical data are available on request from the corresponding author; these data are not publicly available due to privacy or ethical restrictions.

